# Humidity and deposition solution play a critical role in virus inactivation by heat treatment on N95 respirators

**DOI:** 10.1101/2020.06.22.20137448

**Authors:** Nicole Rockey, Peter J. Arts, Lucinda Li, Katherine R. Harrison, Kathryn Langenfeld, William J. Fitzsimmons, Adam S. Lauring, Nancy G. Love, Keith S. Kaye, Lutgarde Raskin, William W. Roberts, Bridget Hegarty, Krista R. Wigginton

## Abstract

Supply shortages of N95 respirators during the coronavirus disease 2019 (COVID-19) pandemic have motivated institutions to develop feasible and effective N95 respirator reuse strategies. In particular, heat decontamination is a treatment method that scales well and can be implemented in settings with variable or limited resources. Prior studies using multiple inactivation methods, however, have often focused on a single virus under narrowly defined conditions, making it difficult to develop guiding principles for inactivating emerging or difficult-to-culture viruses. We systematically explored how temperature, humidity, and virus deposition solutions impact the inactivation of viruses deposited and dried on N95 respirator coupons. We exposed four virus surrogates across a range of structures and phylogenies, including two bacteriophages (MS2 and phi6), a mouse coronavirus (murine hepatitis virus, MHV), and a recombinant human influenza A virus subtype H3N2 (IAV), to heat treatment for 30 minutes in multiple deposition solutions across several temperatures and relative humidities (RH). We observed that elevated RH was essential for effective heat inactivation of all four viruses tested. For heat treatments between 72°C and 82°C, RH greater than 50% resulted in > 6-log_10_ inactivation of bacteriophages and RH greater than 25% resulted in > 3.5-log_10_ inactivation of MHV and IAV. Furthermore, deposition of viruses in host cell culture media greatly enhanced virus inactivation by heat and humidity compared to other deposition solutions such as phosphate buffered saline, phosphate buffered saline with bovine serum albumin, and human saliva. Past and future heat treatment methods or technologies must therefore explicitly account for deposition solutions as a factor that will strongly influence observed virus inactivation rates. Overall, our data set can inform the design and validation of effective heat-based decontamination strategies for N95 respirators and other porous surfaces, especially for emerging or low-titer viruses that may be of immediate public health concern such as SARS-CoV-2.

**Importance:** Shortages of personal protective equipment, including N95 respirators, during the coronavirus disease 2019 (COVID-19) pandemic have highlighted the need to develop effective decontamination strategies for their reuse. This is particularly important in healthcare settings for reducing exposure to respiratory viruses, like severe acute respiratory syndrome coronavirus 2 (SARS-CoV-2), the virus that causes COVID-19. Although several treatment methods are available, a widely accessible strategy will be necessary to combat shortages on a global scale. We demonstrate that the combination of heat and humidity inactivates viruses similar in structure to SARS-CoV-2, namely MS2, phi6, influenza A virus, and mouse coronavirus, after deposition on N95 respirators, and achieves the United States Food and Drug Administration guidelines to validate N95 respirator decontamination technologies. We further demonstrate that depositing viruses onto surfaces when suspended in culture media can greatly enhance observed inactivation, adding caution to how heat and humidity treatments methods are validated.

## Introduction

Effective decontamination of medical equipment is critical for controlling infectious diseases in clinical settings. This is heightened during pandemics, when shortages of personal protective equipment (PPE), such as N95 respirators, lead to occupational risks for healthcare workers. During the coronavirus disease 2019 (COVID-19) pandemic, high demand for N95 respirators has led to interest in decontamination methods that do not compromise the effectiveness of the respirator. The application of heat, UV irradiation, and vaporized hydrogen peroxide are common decontamination treatments in medical settings and research suggests that N95 respirators treated with these methods maintain their filtration integrity and fit (1–4). The United States Food and Drug Administration (USFDA) has issued an enforcement policy for face masks and respirators that presents specific recommendations for validation of PPE decontamination during the COVID-19 crisis. This policy calls for greater than 3-log_10_ inactivation validated using multiple viral pathogens, preferably coronaviruses (e.g., SARS, MERS, MHV, TGEV), and 6-log_10_ inactivation of mycobacteria or bacterial spores (5).

Arguably the simplest and most accessible approach to N95 respirator decontamination is to harness the biocidal activity of heat and moisture. Treating medical equipment with pressurized saturated steam in autoclaves for one hour, for example, leads to high levels of virus inactivation (6–9); however, the high temperatures and pressures in autoclave sterilizers affect N95 respirator integrity (10, 11). In contrast, N95 respirator treatments at lower temperatures, up to 80-90°C for an hour or longer, do not affect filter performance and fit (12–14). To date, studies assessing virus inactivation on N95 respirators at elevated temperatures below 100 °C have included limited viruses and conditions. Influenza viruses heated to 65°C with 85% relative humidity (RH) for 20-30 minutes resulted in greater than 3-log_10_ inactivation (15, 16). Dry heat at 70°C for 1 hour led to > 3-log_10_ inactivation of severe acute respiratory syndrome coronavirus 2 (SARS-CoV-2) (2). In cases where heat treatment is not a feasible decontamination approach, the CDC recommends limited reuse of N95 respirators after incubation in paper bags at room temperature for an excess of five days (17). The justification for this recommendation is based on experiments evaluating stability of SARS-CoV-2 on surfaces (18); however, the efficacy of this practice on N95 respirators has yet to be validated.

Previous virus inactivation studies that focus on other types of surfaces can inform N95 respirator decontamination strategies. Such studies suggest that both temperature and humidity can affect virus inactivation in droplets dried on porous or nonporous surfaces (19). Most surface inactivation studies have focused on virus inactivation at a limited range of environmentally relevant conditions (4 - 40°C) (20–22), and have reported greater virus inactivation at higher temperatures for MS2 bacteriophage (19, 23), enteric viruses (19, 23), coronaviruses (22, 24), and influenza A virus (25). Trends in humidity are not as easily discerned. While some surface inactivation studies have shown increased inactivation at elevated RH (19, 22, 24, 25), others reported that certain nonenveloped viruses are more stable at higher RHs (25). Limited work at elevated temperatures (55 to 65°C) has shown increased influenza virus inactivation as temperature and RH increase (26). A more systematic understanding of how humidity impacts inactivation for a range of viruses would aid the design of effective heat-based decontamination methods.

Cell culture media suspensions are commonly used to generate droplets and aerosols to understand heat inactivation of viruses deposited on surfaces (18, 27, 28). Studies on virus inactivation at room temperature, however, have found that laboratory-made solutions (e.g., artificial saliva, cell culture media) are not representative of respiratory droplets and aerosols (25, 29–31). Indeed, deposition solution composition does appear to impact virus inactivation when virus particles are dried on surfaces, are present in droplets, or are present in aerosols (25, 30–34). Benbough (1971) and Yang et al. (2012), for example, observed that higher protein and salt concentrations positively influenced the persistence of viruses in aerosols and droplets under certain temperature and humidity conditions (30, 32). Protein content also had a protective effect for influenza viruses at median RHs (32). These studies have primarily focused on ambient conditions and the potential impacts of deposition solution on virus inactivation at elevated temperatures has not yet been explored.

An improved understanding of how humidity, temperature, and deposition solution impact virus inactivation in dried droplets is critical to inform PPE decontamination and reuse practices in hospitals and other relevant settings. To address this, we studied virus decontamination on N95 respirator coupons using elevated heat and variable RH for several RNA viruses deposited in droplets from four different solutions. We studied two diverse bacteriophages, MS2 and phi6, a mouse coronavirus (i.e., MHV) and a subtype H3N2 influenza virus (i.e., IAV; see Table S1 in the supplemental material). We focused on RNA viruses because of their relevance to the current COVID-19 pandemic and because of their relevance for other respiratory viruses in clinical settings. Bacteriophage MS2, MHV A59, and IAV are single-stranded RNA viruses like SARS-CoV-2, whereas phi6 is a double-stranded RNA virus. Similar to SARS-CoV-2, phi6, MHV, and IAV are enveloped viruses. We included the two bacteriophages for several reasons, most importantly because their high stock concentrations facilitate experiments with large dynamic ranges. Additionally, the bacteriophages are BSL1 organisms, quickly enumerated, and used extensively in surface decontamination studies, thus allowing for cross-study comparisons. We included an influenza virus, because it is an important human respiratory virus and because there are previous reports of influenza virus inactivation through N95 respirator decontamination processes (15, 16, 35, 36). The mouse coronavirus MHV is in the same genus as SARS-CoV-2 and is thus expected to exhibit a similar fate outside of its host compared to SARS-CoV-2.

Our findings identify key parameters that drive virus inactivation on N95 respirator surfaces with heat and highlight the importance of deposition solution characteristics when validating decontamination methods. In particular, our results suggest that the common practice of depositing viruses using culture media may lead to a significant overestimation of the effectiveness of heat treatment for virus inactivation.

## Results

### Virus inactivation improves with increasing RH and temperature

To understand how temperatures and RHs impact inactivation through heat treatment, we deposited the four RNA viruses in culture media on N95 respirator coupons, and treated the coupons for 30 minutes at 72°C and 82°C. An overall trend of increased virus inactivation was observed as temperature and RH increased (Tables S6 to S9). For all four viruses, inactivation was lowest at 1% RH at both 72°C and 82°C (Figure 1). For treatments with RHs above 25% for 72°C and above 13% for 82°C, inactivation was beyond the detection limits of all viruses. Consequently, we were unable to observe the real inactivation trends at elevated RHs for any of the viruses tested. The dynamic ranges for inactivation varied greatly among the four viruses. Specifically, the dynamic ranges for inactivation were between 5.5-log_10_ and 8.1-log_10_ for the bacteriophages, ∼4.0-log_10_ for IAV, and ∼3.7-log_10_ for MHV.

**Figure 1.**
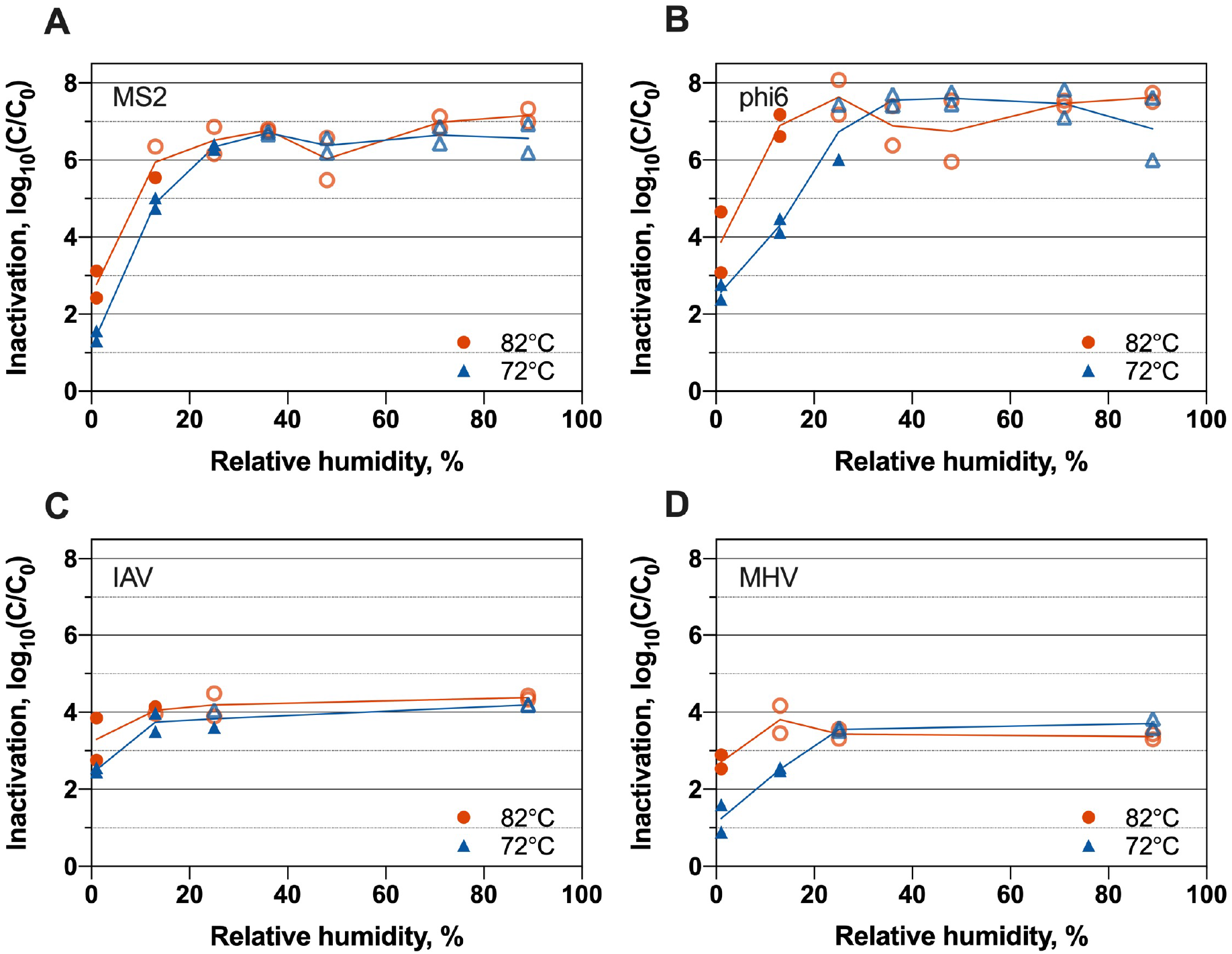
Inactivation of a) MS2 bacteriophage, b) phi6 bacteriophage, c) IAV, and d) MHV at 72°C and 82°C for variable RH when viruses were suspended in culture media. Open faded symbols indicate virus inactivation beyond assay detection limits. Independent experimental replicates (N = 2) are shown for each virus at each condition.

An increase in treatment temperature from 72°C to 82°C resulted in a 1.3-log_10_ and 2.0-log_10_ average increase in inactivation for MS2 and phi6, respectively, across different RHs (Figure 1). This trend of greater inactivation at 82°C compared to 72°C was consistent across nearly all RH conditions for both viruses, though increases were not statistically significant for all settings (Table S10). Likewise, for IAV and MHV at 1% RH, the average inactivation increase was 1.1-log_10_ as temperature increased from 72°C to 82°C (Figure 1). The influence of temperature on inactivation was not calculated for IAV and MHV above 1% RH as assay detection limits were reached.

RH strongly affected virus inactivation. For a given temperature, all four viruses demonstrated increased inactivation with increasing RH for all experiments that fell within the virus assay dynamic ranges (Figure 1). Among the bacteriophages, a 10% increase in RH corresponded to an average 2.5-log_10_ and 2.4-log_10_ increase in inactivation for MS2 and phi6, respectively, across all temperatures within assay limits (Table S11). MS2, for example, underwent an average of 1.4-log_10_ inactivation at 1% RH, and a 4.9-log_10_ inactivation at 13%. MHV and IAV inactivation increased by 1.2-log_10_ and 1.3-log_10_, respectively, from 1% to 13% RH at 72°C.

No consistent trend in inactivation was observed between mammalian viruses and the bacteriophages for treatment conditions within assay limits. For example, the four viruses demonstrated similar inactivation levels at 1% RH at both 72°C and 82°C, but IAV and MHV were inactivated less than the bacteriophages at 72°C and 13% RH (Table S12). The small dynamic range of IAV and MHV limited our ability to determine strong inactivation trends for these viruses.

The MHV and IAV media used in the above experiments consisted of Dulbecco’s modified Eagle’s medium (DMEM) with different supplementary ingredients (Table S4). Control experiments with phi6 and MS2 deposited in both of these media types and treated at 72°C and 13% RH demonstrated that similar inactivation was experienced in either DMEM medium composition (Figure S2).

### Deposition solution influences virus inactivation

To assess potential effects of using tissue culture media as the deposition solution on observed virus inactivation, we conducted phi6 and MS2 experiments over the same range of RHs and temperatures in a PBS deposition solution. We did not include MHV and IAV in these experiments due to the challenges of resuspending MHV and IAV in PBS without decreasing stock concentrations and thus decreasing the experiment dynamic ranges. For both phages, the deposition solution had a profound effect on inactivation (Figure 2). Bacteriophages deposited in IAV medium were inactivated significantly more compared to when deposited in PBS in all conditions for MS2 and three out of four conditions for phi6 (Table S13). The most striking difference occurred at 25% RH for 72°C treatments. At 72°C and 25% RH, for example, only 1.4-log_10_ and 2.9-log_10_ inactivation were observed for MS2 and phi6, respectively, when deposited in PBS. When the bacteriophages were deposited in IAV medium and treated under the same conditions, the observed inactivations were 6.4-log_10_ and > 6.7-log_10_. This is approximately a 4 to 5-log_10_ difference in inactivation resulting solely from type of deposition solution. With either deposition solution, phi6 was more susceptible to heat treatment than MS2. When deposited in PBS solution, phi6 was inactivated, on average, 1.4-log_10_ more than MS2, although significance could not be confirmed for all temperature and RH conditions (Table S14). The difference in inactivation between MS2 and phi6 was less pronounced when viruses were deposited in IAV medium. There, phi6 was inactivated, on average, 0.6-log_10_ more than MS2. We note that differences between phi6 and MS2 inactivation were not due to differences in their respective stock solutions, because the two viruses were deposited in a single mixed stock solution.

**Figure 2.**
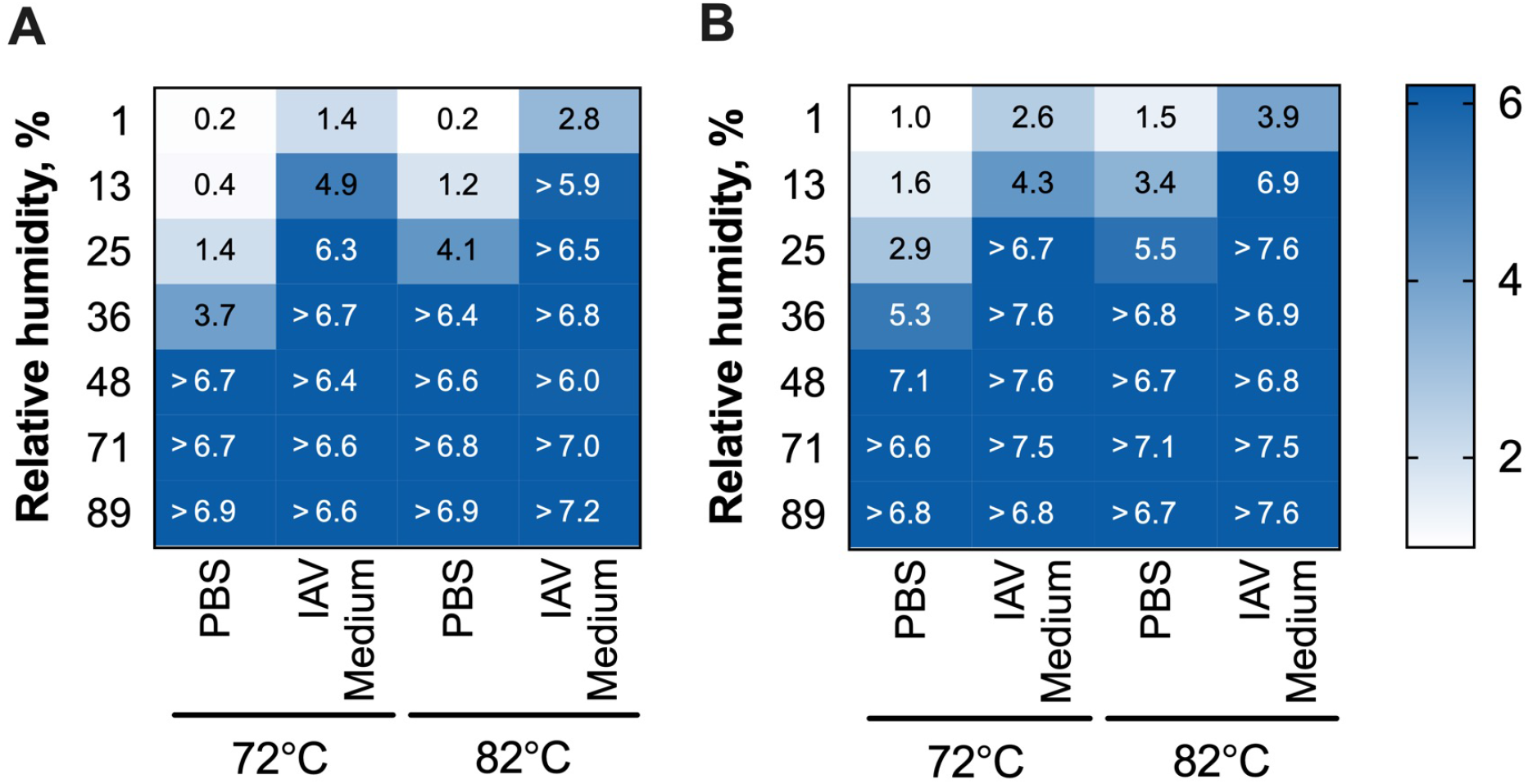
Inactivation of bacteriophages a) MS2 and b) phi6 at variable heat and RH with PBS and IAV medium deposition solutions. Presented values are averages of independent experimental replicates (N = 2) for each condition. Individual replicate data are provided in Tables S6 and S7 in the supplemental material for MS2 and phi6, respectively.

To determine if protein content in IAV contributed to the observed differences in virus inactivation between IAV medium and PBS, we tested MS2 and phi6 inactivation with a PBS deposition solution that was supplemented with the same amount of BSA as was present in the IAV medium. Bacteriophage inactivation in PBS + BSA deposition solution was significantly less than in IAV medium for both MS2 and phi6 across nearly all conditions tested (Figure 3, Table S13), with average reductions of 3.0-log_10_ and 3.7-log_10_ for MS2 and phi6, respectively. With respect to the specific effect of BSA in the deposition solution, MS2 was inactivated 1.3-log_10_ more when the PBS deposition solution contained BSA. Two out of four of the temperature/RH conditions exhibited statistically significant differences (Table S13). For phi6, the opposite trend was observed; adding BSA to the PBS deposition solution resulted in an average of 1.0-log_10_ less inactivation. Here, only one of four conditions tested resulted in a statistically significant difference (Table S13). Overall, these results showed that virus inactivation in PBS + BSA was more similar to inactivation in PBS than in IAV medium for phi6 and MS2.

**Figure 3.**
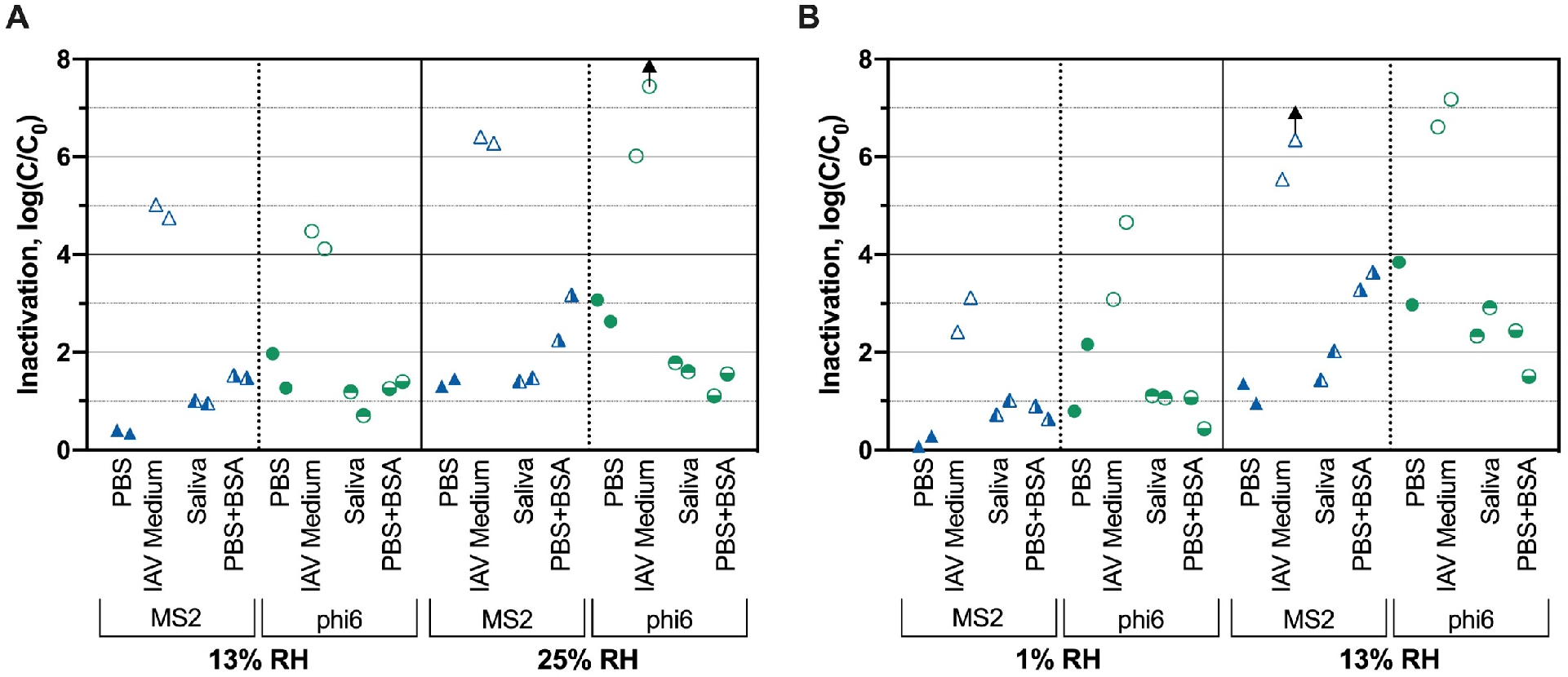
Susceptibility of MS2 and phi6 to heat and RH treatment at a) 72°C and b) 82°C when deposited in four matrices. Arrows indicate virus inactivation beyond detection limits. Independent experimental replicates (N = 2) shown for each virus at each condition.

To better represent virus-containing droplets present on N95 respirators, we tested MS2 and phi6 inactivation in freshly collected human saliva sterilized using UV treatment. Phage inactivation in saliva was within 0.7 log_10_ of inactivation in PBS, on average (Figure 3). Inactivation of the two viruses deposited in IAV medium was significantly greater than inactivation when deposited in saliva across nearly all treatments (3.6-log_10_ and 3.5-log_10_ greater for MS2 and phi6, respectively; Table S13). On average, inactivation levels of MS2 deposited in saliva were 0.5-log_10_ larger than inactivation levels in PBS (Figure 3). For phi6, inactivation in saliva was 0.8-log_10_ less than in PBS (Figure 3). These results indicate that deposition in saliva is more similar to deposition in PBS than in IAV medium.

### Virus inactivation at ambient conditions

To assess whether virus inactivation at ambient conditions was also affected by deposition solution, we tested bacteriophage inactivation at room temperature (20°C) and 36% RH using different deposition matrices. This RH and temperature were selected at the lower end of standard thermohygrometric ranges for healthcare facilities (37). After 24 hours, we observed relatively low levels of inactivation (< 2 log_10_ on average) for both MS2 and phi6 in all deposition matrices (Figure 4). MS2 inactivation in IAV medium was significantly higher than in either PBS (p = 0.0061) or saliva (p = 0.029). Although the average inactivation was higher for phi6 deposited in IAV medium than phi6 in PBS and saliva, the differences were not statistically significant.

**Figure 4.**
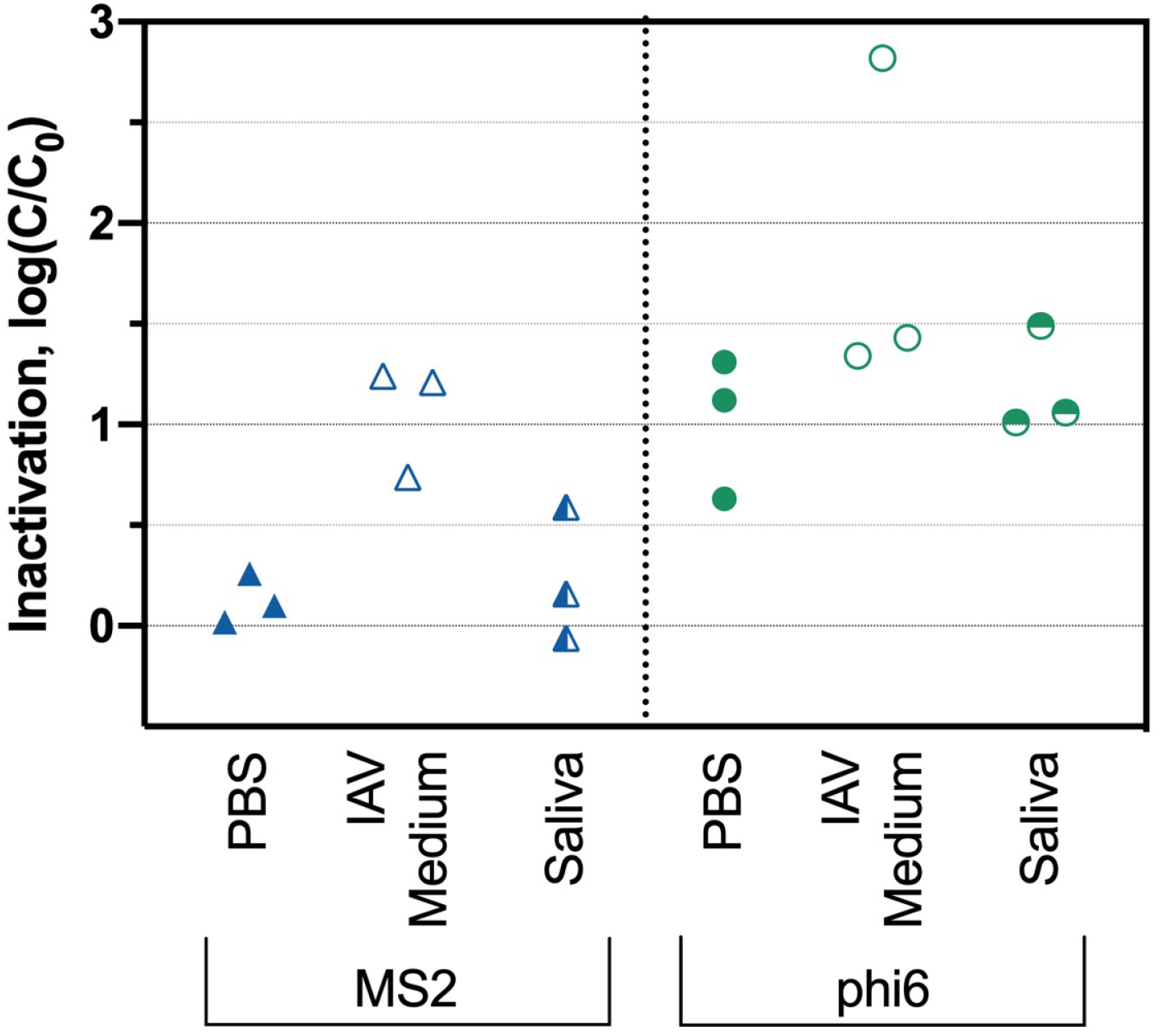
Inactivation of MS2 and phi6 after 24 hours at 20°C and 36% RH. Independent experimental replicates (N = 3) are shown for each virus in each deposition solution.

## Discussion

Heat remains a widely accessible strategy for decontamination of PPE due to the ubiquity of ovens that can achieve sufficient temperatures and the ease of translating and communicating effective protocols. Furthermore, the temperatures and treatment times used in this study do not negatively impact respirator integrity (2, 16, 38), which is essential for this method’s applicability. To support the use of effective N95 decontamination approaches during the COVID-19 pandemic, the USFDA provided a number of emergency use authorizations (EUAs) and published recommendations for evaluating N95 decontamination methods. An early recommendation published by the USFDA for the decontamination and reuse of respirators for single users suggested demonstrating ≥ 3-log_10_ inactivation of viruses, specifically those related to SARS-CoV2 (e.g., SARS-CoV, MERS-CoV, and TGEV), along with ≥ 6-log_10_ inactivation of either mycobacterium or bacterial spores. This guidance was replaced with recommendations to demonstrate greater than or equal to ≥ 6-log_10_ inactivation of three non-enveloped viruses and ≥ 6-log_10_ inactivation of two Gram-positive bacteria and two Gram-negative bacteria (39). The latest guidance does not address which viruses should be tested or which deposition solutions should be used. In addition to identifying the best conditions for heat and humidity decontamination of N95 respirators, our work sought to identify how different surrogate viruses and deposition solutions might affect results.

### Influence of elevated temperature and humidity on virus inactivation

At first glance, our results suggest that increasing temperature improves virus inactivation. When temperature was increased from 72°C to 82°C at a constant RH, an average 1.2-log_10_, 1.6-log_10_, 0.8-log_10_, and 1.5-log_10_ increase in inactivation was observed for MS2, phi6, IAV, and MHV, respectively. Other studies have observed improved virus inactivation at increasing temperatures, both at elevated temperatures (i.e., 55°C vs. 65°C) (26); and at ambient temperatures, ranging from 4 to 40°C (19, 22, 23). Interestingly, when our virus inactivation results are presented as a function of absolute humidity, the effect of increased temperature on inactivation is no longer evident (Figure S3). In other words, the improved inactivation observed for 82°C compared to 72°C was driven more by the added water content than by the temperature. In several studies that have taken a modeling approach, absolute humidity was deemed a better predictor of IAV inactivation than temperature and RH at both elevated temperatures (26) and ambient temperature ranges (40, 41). Prussin et al., however, reported that the conclusions on AH versus RH can vary depending on the type of model used (42). Overall, a mechanistic understanding of what drives virus inactivation on surfaces and in aerosols at various temperatures and humidities remains elusive. Due to the fact that most studies on the effects of humidity on virus inactivation report their findings as a function of RH (19, 22, 23, 25, 27, 32, 43, 44), we chose to present humidities primarily as RH for this study.

Virus inactivation improved as RH increased from 1% to 48% in our experiments at 72°C and 82°C (detection limits were exceeded at all conditions above 48%). A similar RH effect was observed previously for elevated heat treatment of IAV deposited on steel coupons (26). The impact of RH on virus inactivation at lower temperatures is not consistent in the literature. In several studies where diverse viruses have been dried on surfaces and exposed to 4-40°C, increasing RH from 20-30% to 50-80% also increased inactivation (19, 22, 25). Other studies have observed the opposite trend when viruses were dried on surfaces (25) or were present in aerosols (43, 44), with less inactivation at high RH compared to low RH. Phi6 and IAV inactivation at ambient temperatures in droplets, exhibited yet a different trend, with greatest inactivation at RH levels ranging from 60-85% and decreasing at lower and higher RH (30, 42, 45). The reasons for these discrepancies are not clear but may be in part due to differences in the deposition solutions and drying conditions. We suggest that future studies on the impact of temperature and humidity always include a virus that is simple to measure and widely accessible (e.g., bacteriophage MS2) in addition to their viruses of interest (e.g., SARS-CoV-2). Doing so would facilitate cross-study comparisons of various viruses and conditions.

### Virus inactivation at ambient conditions

At room temperature and 40% RH for 24 hours, < 2-log_10_ inactivation of MS2 and phi6 bacteriophages on N95 respirators was observed, regardless of the deposition solution used. Other studies observed less than or equal to 1-log_10_ inactivation of MS2 and phi6 at ambient temperature and RH over 24 hours (27, 46). We observed an additional 0.9-log_10_ inactivation in phi6 compared with MS2, although this difference was not statistically significant. Our tests did not assess the impact of time on ambient condition treatment; we therefore cannot predict the inactivation levels that would be reached when N95 respirators are left at ambient conditions for five days, as specified by CDC guidelines (17). However, previous studies have assessed the persistence of MS2 (27), phi6 (46), and various influenza strains (47), on surfaces over extended time periods. If the rates of inactivation from these studies hold true, 5.5-log_10_ and 4.8-log_10_ inactivation of phi6 and MS2, respectively, on N95 respirators, and 13.6-log_10_ of IAV inactivation on porous surfaces (47), can be expected after five days at ambient temperatures and 30 - 60% RH. Research observing surface stability of clinically relevant viruses at ambient conditions, including IAV and SARS-CoV-2, suggests coronaviruses are not as persistent as IAV or the bacteriophages studied here (18, 35). More work is needed with multiple viruses, longer storage times, and in saliva or other respiratory fluids to determine if room temperature storage is an effective decontamination strategy for N95 respirators.

### Virus-specific inactivation trends

The results of this study demonstrate the importance of assessing inactivation for a diverse set of surrogate viruses when inactivation experiments cannot be performed with the virus of interest because it is not culturable or requires high biosafety containment. In our study, we focused most on the enveloped phi6 bacteriophage and nonenveloped MS2 bacteriophage. To test viruses more similar to SARS-CoV-2, we also included IAV and MHV in a subset of experiments; however, these viruses, like SARS-CoV-2, are difficult to propagate to high titers, thus limiting the experimental dynamic ranges and deposition solutions. Our broad dataset for MS2 and phi6 under different heat/humidity conditions (Figure 2) and in different matrices (Figure 4) suggest that the enveloped phi6 deposited on N95 respirators is more susceptible to heat and humidity than the nonenveloped MS2 under nearly all conditions tested. Previous research on viruses dried on surfaces and exposed to room temperature also found increased persistence of non-enveloped viruses compared to enveloped viruses (48, 49). As viruses dry on surfaces, it has been suggested that the air water interface may damage the lipid membranes of enveloped viruses (19, 30) and the increased salt concentration can cause the lipid membrane to become rigid (50). Moreover, some studies observed that increased salt concentrations can protect non-enveloped viruses (30, 32).

We did not observe that the enveloped mammalian IAV and MHV viruses were consistently more susceptible to inactivation than the non-enveloped MS2 (Figure 1). In fact, MHV was less susceptible than both bacteriophages to heat treatment at 72C and 13% RH when deposited in culture media (Figure 1). We note that our dynamic ranges for MHV and IAV were much smaller than those for MS2 and phi6. Furthermore, MHV and IAV experiments were only conducted when using their culture media as deposition solutions. These limitations of the IAV and MHV experiments affect our ability to observe major trends in their relative susceptibilities. Nonetheless, the results from experiments with all four viruses suggest that bacteriophages are not always conservative surrogates for IAV and coronavirus inactivation through heat and humidity treatment. This brings to question the USFDA’s guidelines for using non-enveloped viruses as conservative surrogates for pathogenic viruses on N95 respirator decontamination. A review by Yang and Marr on virus survival in aerosols indicates that the presence of a lipid envelope is not solely responsible for virus susceptibility to inactivation; they suggest other virus characteristics, such as virus infection mechanisms and protein stability, are necessary to explain observed inactivation levels (51). To better account for these differences, a “cocktail” approach to assessing virus inactivation may be most suitable, using a wide range of surrogate viruses that have various characteristics in common with the viral pathogens of interest.

### Effects of deposition solution

Our results demonstrate that the deposition solution used to apply viruses to N95 respirators greatly impacts virus inactivation, both at elevated and ambient temperatures. At elevated temperatures, both MS2 and phi6 were inactivated much more when deposited in their culture media relative to PBS (Figures 2 and 3). At room temperature, only MS2 was inactivated to a greater extent when deposited in its culture medium (Figure 4). There has been limited prior work comparing virus survival in PBS versus culture media. In contrast to our results, Yang et al observed that IAV deposited in culture media supplemented with fetal calf serum exhibited less inactivation compared to viruses deposited in PBS when exposed to room temperature and RH from 20 to 60% (30). This discrepancy may be due to the fact that the deposition solution was allowed to dry before heat treatment in our study, whereas Yang et al. measured inactivation while viruses were still suspended in droplets. Consistent with this hypothesis, Sizun et al observed human coronaviruses OC43 and 229E in suspension exhibited less inactivation in PBS than in culture media with fetal bovine serum at room temperature (21).

There are multiple possibilities that may explain the observed differences in inactivation between PBS and culture media deposition solutions. Studies in aerosols or droplets have suggested increasing salt concentrations in media may increase inactivation (30, 32); however, the PBS and IAV medium solutions in our study had similar salt content (Tables S2 and S3). Proteins in the deposition solution could have a protective effect on viruses (29, 30). Our results, however, show that inactivation with viruses deposited in PBS + BSA was significantly less than when applied in IAV medium, which contains BSA at the same levels (Figure 4). Therefore, protein content also does not appear to explain the observed differences. Another possible explanation is that the L-glutamine present in IAV medium degrades into glutamate and ammonia (52, 53). Ammonia is known to cause virus inactivation in solution (54), although further work is needed to test this hypothesis in dried droplets and at elevated temperatures.

Ultimately, it is important to understand the extent of inactivation when viruses are deposited in the actual matrices found on N95 respirators. The limited studies that have compared artificial and more realistic deposition matrices (e.g. human saliva) suggest that laboratory made solutions (e.g. cell culture media or artificial saliva) are not fully representative of respiratory droplets and aerosols (29, 34, 35, 55). For this reason, we tested human saliva as a more realistic deposition solution and compared the results to those obtained with the other deposition solutions. Our results show that MS2 and phi6 were significantly more susceptible to inactivation in IAV medium as opposed to saliva (Figure 4). Furthermore, the PBS deposition solution provided the most similar results to the saliva deposition solution. These results suggest that PBS may be an appropriate deposition solution for studying virus inactivation on surfaces. Additional experiments will need to be conducted with a broader set of viruses to determine if a PBS deposition solution is always representative of saliva.

In light of our results, it is important to consider the deposition solution when re-examining earlier reports of virus inactivation on N95 respirators and other materials. Heat inactivation studies and room temperature inactivation studies often either use culture media to generate droplets and aerosols or do not explicitly state which solutions are used (2, 15, 16, 18, 27, 28). Given that our observed MS2 and Phi6 inactivation trends hold for other viruses, then inactivation in several studies using culture media for droplet deposition may overestimate inactivation relative to what occurs when viruses are dried on N95 respirators in saliva or respiratory fluids. A recent study of SARS-CoV-2 inactivation on N95 respirators, for example, reported 3-log_10_ virus inactivation at 70°C under “dry heat” conditions for 60 minutes (2). Although the deposition solution was not explicitly stated in this study, inactivation of SARS-CoV-2 could have been significantly overestimated if a culture medium was used for deposition. This is of critical importance for clinical settings, because these results may lead healthcare workers to disinfect respirators at 70°C for 60 minutes in an oven without controlled humidity whereas our results at 72°C and 1% RH for 30 minutes suggest little inactivation would take place under these conditions.

## Conclusions

Our work demonstrates the virus inactivation efficacy of heat and humidity treatments for N95 respirator decontamination. The USFDA’s recommended 6-log_10_ inactivation of viruses was easily achievable for bacteriophages MS2 and phi6 with this heat-humidity paradigm. Likewise, although we were limited by the dynamic range of our assays, the more clinically relevant virus surrogates, MHV and IAV, resulted in at least 3-log_10_ inactivation under the same conditions. Low (< 25%) RH treatments at the same temperatures were not as effective. We also observed that inactivation was strongly influenced by the deposition solution. Dried virus droplets in cell culture media were inactivated significantly more than in any other deposition solution (PBS, PBS + BSA, saliva). These findings suggest that virus inactivation may be vastly overestimated when using culture media as the deposition solution in surface disinfection studies. We suggest the use of deposition solutions more similar to human saliva or respiratory fluid in virus inactivation experiments to ensure representative results.

Hospitals and other healthcare settings can expect extensive virus inactivation of N95 respirators through heat treatment for at least 30 minutes at 72°C or 82°C and RH above 50%. High humidity heat treatment is particularly appealing as it can be readily adapted and scaled to a range of settings, from health care facilities to private residences. Further, implementation is equally suitable for healthcare systems or individuals without access to specialized equipment, including those in low-to middle-income countries. These results provide timely and useful information for efficacious N95 respirator decontamination, enabling reuse when necessary due to shortages.

## Materials and Methods

### Virus stocks and enumeration

MS2 bacteriophage and its *Escherichia coli* host were obtained from the American Type Culture Collection (ATCC 15597). Bacteriophage phi6 and its *Pseudomonas syringae* pv. phaseolicola host were provided by Linsey Marr at Virginia Tech. MHV strain A59 and its murine delayed brain tumor (DBT) host cell line were provided by Julian Leibowitz at Texas A&M Health Science Center College of Medicine. For IAV, we used a recombinant virus that expresses the luciferase reporter in infected cells. This system allows for rapid titering based on light emission in infected cells. The virus is a 6+2 reassortant, in which the genomic segments encoding the surface hemagglutinin (HA) and neuraminidase (NA) are derived from A/Wisconsin/67/2005 (H3N2) and the remaining six segments are derived from A/WSN33 (H1N1). In this case, the segment 3 RNA encodes a polymerase acidic (PA) protein that is fused to the NanoLuc reporter (PMCID: PMC3838222). Recombinant viruses were harvested after transfection of HEK 293T/MDCK-SIAT1 co-cultures with plasmids expressing the genomic RNA and proteins of all 8 segments. Rescued viruses were passed once on MDCK-SIAT1 cells at an MOI of 0.05 to obtain a passage 1 (P1) stock.

### Virus propagation and purification

The MS2 viruses were propagated and titered based on established methods (56, 57). The MS2 lysate was concentrated with polyethylene glycol 8000 (Fisher Scientific, Cat. No. BP2331), treated with chloroform, and filter-sterilized with a 0.22 µm polyethersulfone (PES) membrane filter (CELLTREAT Scientific, Cat. No. 229746). Propagated phi6 was filtered with a 0.22 μm PES membrane filter, concentrated by tangential flow filtration (Millipore, Cat No. C1975) with a 30 kDa cellulose filter (Millipore, Cat. No. PXC030C50), purified by sucrose gradient ultracentrifugation, and filtered-sterilized with 0.22 μm PES membranes (58). The MS2 and phi6 experimental stocks were combined, resulting in a single bacteriophage stock with each virus present at ∼1011 PFU/ml. The stocks were aliquoted and stored at -80°C prior to use.

For MHV propagation and enumeration, DBT cells were grown in Dulbecco’s modified Eagle’s medium (DMEM; Lonza, Cat. No. 12614F) supplemented with 10% horse serum (Life Technologies, Cat. No. 26050088), 1% penicillin streptomycin (Invitrogen, Cat. No. 15140122), and 1% L-glutamine (Invitrogen, Cat. No. 25030081) at 37C and 5% CO2. Cells were infected at a multiplicity of ∼0.01 when they were 75% confluent, then incubated with virus for 24 hours. The cell suspension was frozen at -80°C, thawed, centrifuged at 3,000 x g for 15 min at 4°C, and then the supernatant was recovered. The resulting virus stock (∼106 PFU/ml) was filtered with a 0.22 µm PES membrane and stored in single-use aliquots at -80°C. For plaque assay enumeration, samples were diluted in MHV medium (DMEM with 2% horse serum, 1% penicillin and streptomycin, and 1% L-glutamine) and applied to confluent DBT cells washed with 1X PBS (Invitrogen, Cat. No. 10010023). After one hour of incubation at room temperature with rocking, the virus suspension was removed from monolayers and an overlay of 1.6% agar mixed 1:1 with a 2x Minimum Essential Medium (MEM; Quality Biological, Cat. No. 115073101) containing 5% horse serum, 10 mM HEPES (Lonza, Cat. No. 17737E), 1X MEM Non-essential amino acids (Invitrogen, Cat. No. 11140050), 2% L-glutamine, and 2% penicillin streptomycin were applied. Infected cells were then incubated for 48 hours at 37°C and 5% CO2 before staining with neutral red solution (Sigma-Aldrich, Cat. No. N2889) diluted in 1X PBS to a 0.01% final concentration. Plaque assays were conducted in triplicate and a negative medium control was performed with each assay.

IAV viral propagation was performed in IAV medium (DMEM (Gibco, Cat. No. 11965092) with 25 mM HEPES pH 7.2-7.5, 0.1875% Fraction V bovine serum albumin (BSA; Gibco, Cat. No. 15260037), 1% penicillin and streptomycin (10,000 U/mL; Gibco, Cat. No. 15140122), and 2 µg/ml TPCK-Treated Trypsin (Worthington Biochemical Corporation, Cat. No. LS003740). IAV stocks were stored as single-use aliquots in 0.5% glycerol at -80°C. IAV was enumerated in MDCK-SIAT1 cells by endpoint dilution using IAV titer media, which contained 1% BSA, but otherwise had the same components as IAV medium. Eighteen hours post-infection, media were aspirated and replaced with IAV titer media containing 7.5 µM ViviRen Live Cell Substrate (Promega, Cat. No. E6491). Light emission was measured using a BioTek Synergy HTX luminometer using the following settings: 3-minute dark adapting hold, Emission-Hole, Optics Position-Top, GAIN 160, Integration Time-1.00 seconds, Read Height-2.24 mm, room temp. A well was considered positive for infection if the RLU were greater than or equal to twice the average background RLU from eight mock-infected wells.

### Droplet deposition

The combined bacteriophage stock was suspended to a final concentration of approximately 10^10^ PFU/ml in various deposition solutions. These deposition solutions (Table S2), each used in a subset of experiments, included 1X PBS (Table S3), IAV medium, with the exception that no trypsin was added (Table S4), MHV medium (Table S4), PBS with 0.1875% BSA (PBS + BSA), or human saliva. For each saliva experiment, fresh saliva was collected from a volunteer and UV treated to sterilize (59, 60). Volunteers did not eat within 2 hours prior to collection and rinsed their mouth with water ten minutes before collection (61, 62). Saliva was collected in two wells of a 12-well plate in a thin layer. The saliva was immediately treated for five minutes using a custom-built collimated beam equipped with 0.16 mW cm^-2^ UV_254_ lamps (model G15T8, Philips). Lamp intensity was measured using chemical actinometry (63, 64). MHV (∼10^6^ PFU/ml) in MHV media (Table S4) and IAV (∼10^7^ TCID_50_/ml) in IAV media (Table S4) were used for droplet deposition.

A total of 50 μL of the virus suspensions were deposited in 25 2 µL droplets on 2.54 cm diameter circular coupons. The coupons were generated from 3M 1860 N95 respirators with a 2.54 cm arch punch. The droplets were then allowed to dry in a biosafety cabinet at room temperature and ambient RH for approximately one hour. Details of the experiment are described in the supplemental material. Each treated coupon had a corresponding control coupon that was prepared at the same time as the treated coupon but was maintained at ambient conditions during the experiments.

### Temperature and humidity controlled oven

The temperature and humidity controlled oven (TestEquity 123H Temperature/Humidity Chamber) used in all experiments was calibrated for temperature to be accurate within 2°C and RH to be accurate within 5%. A second external instantaneous hygrometer probe (Fisher Scientific, Cat. No. 116617B) rated to be accurate within 0.2°C and 1.5% RH was also used to monitor the oven. The experiments were designed to test regular intervals of RH using the humidity chamber’s readout (i.e., 10%, 20%, 30%, 40%, 50%, 70%, 90%), however, due to the lower error associated with the external hygrometer, external hygrometer values (1%, 13%, 25%, 36%, 48%, 71%, 89%) were used in the data analysis (Table S5).

### Heat and RH controlled experiments

The heat and humidity controlled oven was set to the desired RH and temperature for at least 30 minutes prior to use. Dried coupons were immobilized on a coated metal test tube rack with metal binder clips (Figure S1). The metal rack with coupons was then transferred to the oven at the predetermined temperature and humidity settings. Treatment times were started when the oven RH was within 1% of the target RH for 13 - 71% RH and within 5% of target RH for 1% and 89% RH. The oven reached these conditions within five minutes. Heat decontamination experiments with phi6 and MS2 deposited in PBS and IAV media were conducted at 72 °C and 82 °C and 1%, 13%, 25%, 36%, 48%, 71%, 89% RH. Experiments with IAV and MHV were carried out in duplicate for a subset of experimental conditions (i.e., temperatures of 72 °C and 82 °C, each at 1%, 13%, 25%, 48%, and 89% RH). Experiments with additional deposition solutions, including human saliva and PBS with BSA, were carried out for MS2 and phi6 at 72°C with 13% and 25% RH and at 82°C with 1% and 13% RH. The IAV and MHV DMEM media contained different supplementary ingredients; to assess the possible effects of these differences on observed virus inactivation, duplicate control experiments with MS2 and phi6 at 72°C and 13% RH were carried out in both MHV media and IAV media.

Twenty-four hour experiments were conducted with MS2 and phi6 to evaluate virus inactivation that would take place on N95 respirators when stored at ambient conditions in healthcare settings. MS2 and phi6 suspended in IAV medium, PBS, PBS with 0.1785% BSA, and saliva, were deposited on N95 respirator coupons as described above for the elevated temperature experiments. Coupons were then incubated in the temperature and humidity-controlled oven at 20°C and 36% RH for 24 hours. Infective virus concentrations on coupons following 24 hours of incubation were compared to the infective viruses on control coupons that were enumerated immediately after droplets were dried for one hour. Three independent replicates of the experiment were conducted.

### Virus extraction

To recover viruses from control and treated coupons, the coupons were cut into 4-6 pieces with sterilized scissors and suspended in 1.3 ml elution medium. For phi6 and MS2, the elution media consisted of 1.3 mL of 1% BSA (Dot Scientific, Cat. No. DSA30075) in PBS. For MHV, the elution solution was MHV media. For IAV, the elution solution was IAV titer media (same as IAV media, except 1% BSA is used in place of 0.1875% BSA). The coupon suspensions were vortexed at medium speed for 1 min. Viruses extracted in the elution buffer were enumerated as described above. To assess recovery from the coupons, 50 µL of the virus deposition solution was suspended into 1.3 mL of 1X PBS with 1% BSA, MHV media, or IAV titer media for phages, MHV, and IAV, respectively. Virus recovery was determined as the ratio of the control coupon virus titer to the suspended virus solution and were greater than 7% for all viruses and conditions.

### Statistical analyses

Unpaired t-tests were performed to determine differences in virus inactivation for different treatment conditions and viruses using GraphPad Prism 8 software. Statistical significance was considered as p-values < 0.05.

## Data Availability

Data is included in the SI document.

## Acknowledgments

We thank Brittany Hicks for assistance preparing materials and Brad Angelocci, Curt Cooper, Patrick McNally, Ken Arnett, and Bryan Shores for their generous support with the humidity chamber.

This work was supported by the University of Michigan College of Engineering and the University of Michigan Health System. N.R. was supported by a UM Rackham Predoctoral Fellowship, L.L. was supported by NSF INFEWS T3 grant number 1632974, P.A. was supported by the Water Research Foundation grant #15-07, K.H. was supported by Hampton Roads Sanitation District, K.L. was supported by an NSF GRFP fellow ID 2016216003, and B.H. was supported by a UM COE Blue Sky Grant.

